# Aerobic exercise improves executive function after traumatic brain injury via changes to the functional connectivity of the anterior cingulate cortex

**DOI:** 10.64898/2026.02.27.26347275

**Authors:** Emma M. Tinney, Mark C. Nwakamma, Goretti España-Irla, Madeleine L. Perko, Lauren Kong, Colette Chen, Jeremy Hwang, Amanda O’Brien, Ryan Luke Sodemann, Jacqueline Caefer, Julia Manczurowsky, Charles H. Hillman, Alexandra Stillman, Timothy P. Morris

## Abstract

Executive dysfunction affects nearly 50% of individuals with traumatic brain injuries (TBI), yet interventions targeting the underlying neural mechanisms remain limited. This study examined whether aerobic exercise modulates functional connectivity to improve executive function in individuals with mild TBI and identified the neural pathways mediating these improvements. In this secondary analysis of a 12-week pilot randomized controlled trial, participants with mild TBI (n=24) were randomized to aerobic exercise (n=12) or active balance control (n=12). Resting-state fMRI with multivariate pattern analysis revealed that aerobic exercise selectively altered functional connectivity patterns of the anterior cingulate cortex (ACC) compared to balance control. Post-hoc seed-to-voxel analyses identified widespread ACC connectivity differences between groups post-intervention while controlling for baseline, across 19 cortical regions spanning default mode, frontoparietal control, and salience networks. Critically, greater anticorrelation between the ACC and insula following aerobic exercise was associated with improved Trail Making Test B-A performance in the aerobic group (β=46.92, p=0.04) but not the balance group, indicating that participants who developed stronger ACC-insula functional segregation showed greater reductions in executive function completion times. These findings establish the ACC-insula circuit as a critical neural substrate mediating exercise-induced executive function recovery after TBI and identify this pathway as a promising therapeutic target for exercise-based rehabilitation interventions.

## Introduction

Executive dysfunction affects nearly 50% of individuals with traumatic brain injuries (TBI), manifesting as deficits in attention, working memory, and cognitive flexibility that impair activities of daily life, such as return to work, driving, and functional independence(Rabinowitz & Levin, 2014). Despite this substantial burden, no FDA-approved pharmacological treatments exist for cognitive dysfunction following TBI, requiring alternative therapeutic approaches.

Aerobic exercise has emerged as a promising non-pharmaceutical intervention, demonstrating selective improvements in executive functioning across multiple populations(Dupuy et al., 2024). In TBI specifically, studies have shown that aerobic exercise interventions improve executive functioning performance(Chin et al., 2015; López et al., 2024; Morris et al., 2016; Sharma et al., 2020; Snowden et al., 2023; Tinney et al., 2025), with benefits observed across injury severities from mild to severe TBI. However, the neural mechanisms underlying exercise induced cognitive improvements remain unknown, demonstrating a critical gap that limits intervention optimization and evidence-based implementation.

Emerging evidence from the broader exercise neuroscience literature provides a mechanistic framework linking aerobic activity to brain network reorganization. In healthy older adults, aerobic exercise interventions modulate functional brain connectivity within cognitively relevant networks, with connectivity changes predicting executive function gains (Voss et al., 2010). Similarly, other studies suggests that aerobic exercise and higher cardiorespiratory fitness is related to functional connectivity in networks supporting cognition (Moore et al., 2022). Preclinical TBI models further demonstrate that post-injury exercise promotes neurological recovery through neurogenesis(Ho et al., 2009), modulated inflammatory responses(Piao et al., 2013), and improved cerebral blood flow(Zhang et al., 2013). While these cellular mechanisms cannot be measured non-invasively in humans, functional magnetic resonance imaging (fMRI) provides a validated non-invasive proxy for estimating underlying neuronal network activity and connectivity patterns (Mishra et al., 2020). As such, we aimed to investigate the underlying cortical plasticity changes that support executive functioning improvement after aerobic exercise in mild TBI.

Building on our prior behavioral findings showing executive functioning improvements following aerobic exercise in this cohort (Tinney et al., 2025), the current study tested for differences in functional connectivity between intervention arms and whether any differences mediated cognitive improvements. We hypothesized that aerobic exercise, compared to balance exercise, would modulate resting state functional connectivity supporting executive functioning, and that these functional connectivity changes would be associated with the magnitude of executive functioning improvement. Using a data-driven multivariate pattern analysis, we examined differences between groups in resting-state functional connectivity.

## Method

Details of all study protocols can be found in Tinney et al(Tinney et al., 2026) and described briefly below. This study received ethical approval from Northeastern University Institutional Review Board.

### Participants

Adults aged 18-55 years with a diagnosis of mTBI within the last one year were recruited for participation in an exercise intervention (The Exercise and Concussion Health Study (TECHS)) (ClinicalTrials.gov: NCT06494592). Participants were excluded if they had a skull breach, subdural hematoma, prior diagnosis of cognitive or physical disability, clinical diagnosis of neurological or neuropsychiatric disorder, undergoing treatment for cardiovascular events, not fluent in English, not medically cleared for exercise, use of an assisted walking device, or not MRI compatible. All participants received a formal concussion or mTBI diagnosis by a physician (AS). Diagnoses were confirmed using OSU-TBI-ID questionnaire, requiring a direct blow or impact to the head with either a loss of consciousness of <5 minutes, dazed, or had a gap in memory. 37 participants were enrolled and randomized into the exercise intervention. Nine participants dropped out of the study, and four were excluded from final analyses due to completing the endpoint too late (n=2) and being too familiar with cognitive tests (n=2). See Table 1 for participant demographics.

**Table 1:**
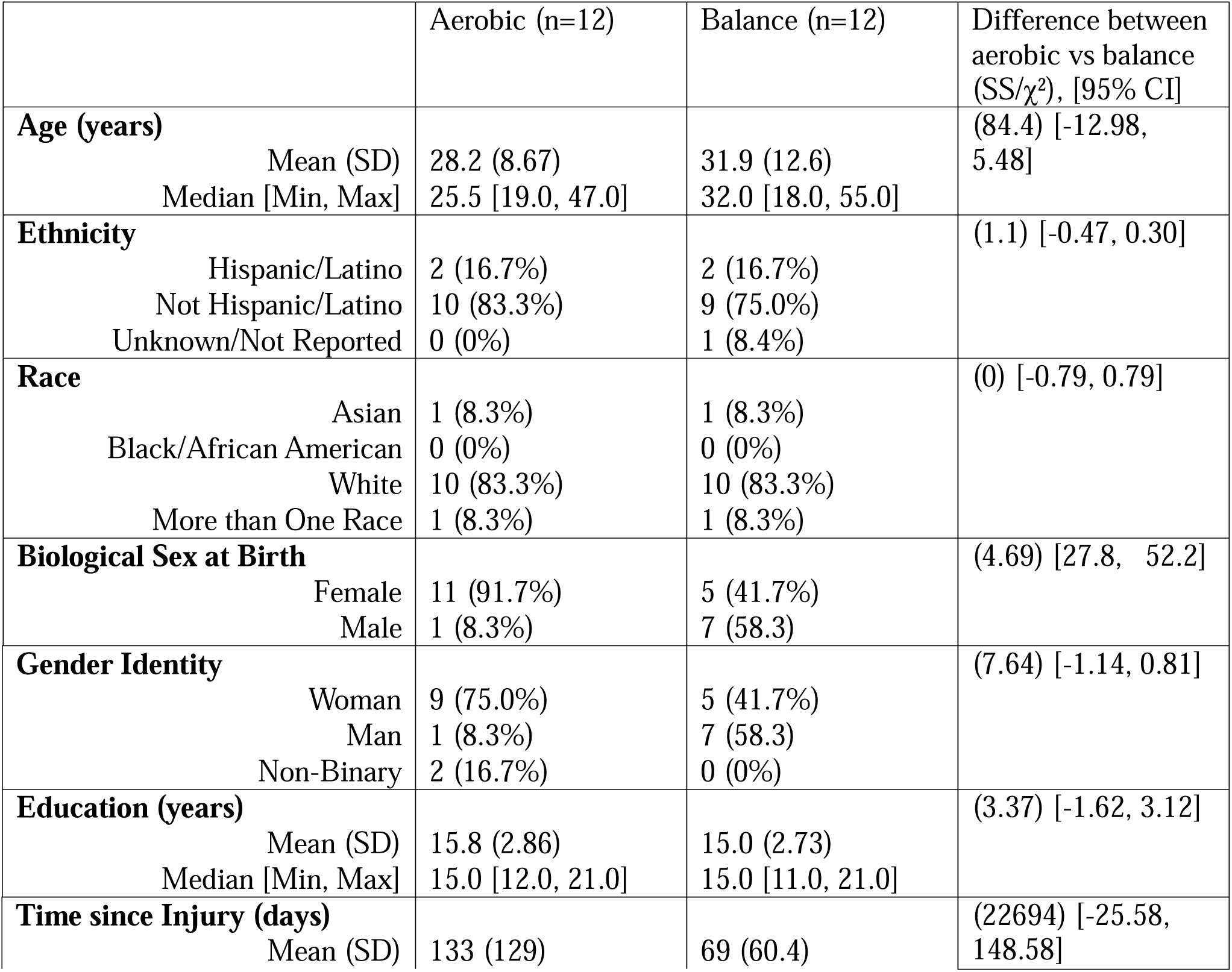
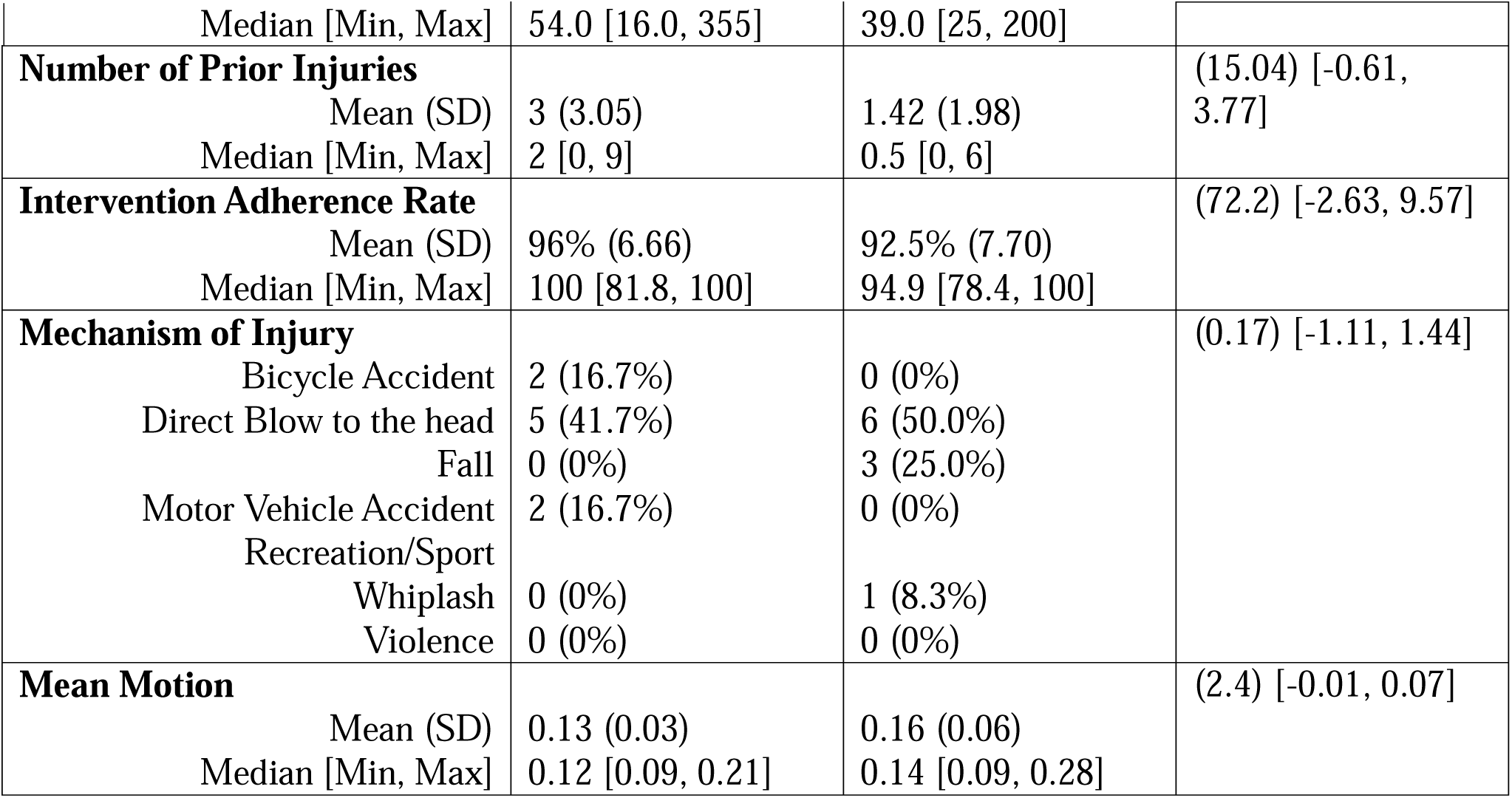
Participant Demographics.

### Design

Participants were randomized after a baseline session using a block randomization in block sizes of four and six with equal allocation to each group in REDcap (n=12 per group). All participants engaged in one-on-one staff-led virtual exercise interventions three times a week for 30-minutes (five-minute warm up, 20-minutes of aerobic/balance exercises, five-minutes cooldown) for 12 weeks, making a total of 36 intervention sessions. Three sets of five exercises, with modifications per each exercise, were developed for each intervention group (balance and aerobic) to help maintain engagement and allow intensity of the exercise to progress (see protocol paper for details). All virtual sessions were administered by two trained research staff members, one who led the exercise session and one who monitored safety. Heart rate (HR) was monitored and recorded by hyperate4health (https://hyperate4health.netlify.app/), a proprietary open access platform and participants were asked to rate their symptoms and exertion every five minutes. Continuous HR recording via hyperate4health did not work for all participants, so heart rate was also recorded every 5 minutes into RedCap. Aerobic sessions had a goal of achieving 20 minutes of aerobic exercise performed at 80% of HR at symptom threshold. If this intensity increased symptoms by +3 on the symptom checklist, a one-minute break was taken, and the goal was reduced to 60% of symptom threshold HR. If this intensity still exacerbated symptoms, another one-minute break was taken, and exercises were done with a goal of 40% of symptom threshold HR.

### Executive function assessment

The trail making test (TMT) was administered before and after the intervention by two trained staff members (ET and MN). Three scores were derived from this test. TMT-A requires participants to connect numbered circles in sequential order (1-2-3…) and primarily measures processing speed, visual scanning, and basic motor function. TMT-B requires participants to alternate between numbers and letters in sequential order (1-A-2-B-3-C…) and assesses executive function, including set-shifting, cognitive flexibility, and working memory, in addition to the processing speed demands present in TMT-A. The TMT B-A difference score isolates executive function demands (set-shifting, cognitive flexibility) by subtracting the processing speed component (TMT-A) from the combined executive function and processing speed demands (TMT-B). Lower scores on all three metrics (TMT-A, TMT-B, and TMT B-A) indicate better performance, as they reflect faster completion times. The B-A difference score is particularly valuable in TBI populations as it controls for individual differences in motor speed and visual processing that might confound interpretation of executive function abilities. A further battery of tests was conducted but not analyzed here as no intervention effects were found in the initial behavioral analysis.

### MRI Data Acquisition

A Siemens Prisma 3T scanner, with a 64-channel head coil was used to complete MRI scans pre and post intervention. Structural imaging consisted of high-resolution three-dimensional (3D) anatomical sequences including a Magnetization-Prepared Rapid Acquisition with Gradient Echo (MPRAGE) T1-weighted sequence with navigator correction (sagittal orientation, 0.8 mm isotropic resolution, TE/TI/TR = 1.81-7.18/1000/2500 ms, field of view (FOV) 256 mm × 240 mm, 208 slices, GRAPPA acceleration factor = 2). An anatomical scout sequence was also acquired for positioning (sagittal orientation, 1.6 mm isotropic resolution, TE/TR = 1.37/3.15 ms, FOV 260 mm, 128 slices, GRAPPA acceleration factor = 3). Functional MRI included a Echo Planar Imaging (EPI) sequence (2.0 mm isotropic resolution, TE/TR = 37/800 ms, multiband acceleration factor = 8, FOV 208 mm, 72 slices, 600 measurements, PA phase encoding direction, total time = 8 minutes and 10 seconds). Field mapping sequences for distortion correction were acquired with matching geometry (2.0 mm isotropic resolution, TE/TR = 66/8000 ms, FOV 208 mm, 72 slices) in both AP and PA phase encoding directions.

### MRI Preprocessing

Images were preprocessed using fmriprep(Esteban et al., 2019), then further preprocessed using CONN(Whitfield-Gabrieli & Nieto-Castanon, 2012) and SPM(Penny et al., 2011). After fmriprep, which included head motion correction, realignment, slice timing correction for sequential acquisition, susceptibility distortion correction, co-registration to reconstructed structural images, and spatial normalization to standard space, functional data was smoothed using spatial convolution with a Gaussian kernel of 8 mm full width half maximum (FWHM). In addition, functional data were denoised using a standard denoising pipeline including the regression of potential confounding effects characterized by white matter timeseries (5 CompCor noise components), CSF timeseries (5 CompCor noise components), motion parameters and their first order derivatives (12 factors)(Friston et al., 1996), outlier scans (below 62 factors(Power et al., 2014), session and task effects and their first order derivatives (4 factors), and linear trends (2 factors) within each functional run, followed by bandpass frequency filtering of the BOLD timeseries(Hallquist et al., 2013) between 0.008 Hz and 0.09 Hz.

CompCor(Behzadi et al., 2007; Chai et al., 2012) noise components within white matter and CSF were estimated by computing the average BOLD signal as well as the largest principal components orthogonal to the BOLD average, motion parameters, and outlier scans within each subject’s eroded segmentation masks. From the number of noise terms included in this denoising strategy, the effective degrees of freedom of the BOLD signal after denoising were estimated to range from 135.3 to 150.1 (average 148) across all subjects.

### Multivariate Pattern Analysis (MVPA)

MVPA is a data-driven connectivity approach used for whole brain voxel wise resting state connectivity analysis, reducing research degrees of freedom. MVPA has demonstrated significant utility in uncovering neural differences in TBI and controls(España-Irla et al., 2025), functional changes in response to exercise(Cline et al., 2024), and fitness related functional connectivity and cognition following an exercise intervention(Lloyd et al., 2024). MVPA analyzes functional connectivity by examining each voxel within the brain and calculating its connectivity to all other voxels in the brain for each participant. For any given seed voxel, the connectivity maps from all participants are combined and summarized using a dimensionality reduction technique, with singular value decomposition applied to the connectivity matrix generating an eigen pattern score (analogous to principal component analysis). This eigenpattern score serves as the dependent variable in a group-level general linear model testing the difference in endpoint functional connectivity between groups while controlling for baseline significantly differences from zero at that voxel. This analytical process is systematically applied to every voxel in the brain, producing a whole brain statistical parametric map. We analyzed 64 within-voxel dimensions and extracted the first five factors.

Multiple comparison correction was implemented using nonparametric cluster level inference (voxel level threshold: p <0.001, cluster level threshold: p < 0.05, family wise error corrected) enabling statistical inference about functional connectivity patterns between suprathreshold clusters and the rest of the brain. The resulting connectivity maps represent the relationship between endpoint differences in whole brain resting state functional connectivity between groups. To further characterize these connectivity patterns, suprathreshold clusters were used as seeds in a follow up seed-to-voxel analysis. While these post-hoc analyses contain inherent bias, they provide valuable insights for interpretation and hypothesis generation (Nieto-Castanon, 2022). Detailed methodological specifications are available in Nieto-Castanon (2022). All analyses were conducted using CONN Toolbox with default pipeline settings and standard cluster biased inference procedures based on Gaussian Random Field Theory(Worsley et al., 1996).

### Statistical Analysis

All subsequent statistical analyses were performed in RStudio Version 4.1. To test for association between change in network based functional connectivity and change in TMT performance linear mixed-effects models, with main effects of rs-fMRI metric change score from, and a group*time interaction coupled with a participant-specific random slope were performed separately for TMT B and TMT B-A. To reduce the number of comparisons, we extracted the functional connectivity between the ACC and the average of each suprathreshold cluster within a network). False discovery rate (FDR)-corrected simple slopes were tested post hoc to determine any marginal effects of the interaction term. Education, age, biological sex, days since injury and mean framewise displacement were accounted for in all models.

## Results

As previously reported(Tinney et al., 2025), the aerobic group demonstrated faster competition time on TMT-B (β=13.15,SE= 6.05,g=1.24,95%CI[0.04,2.45]) and TMT B-A (β=13.36,SE=6.31,*g*=1.20,95% CI[0.00,2.41]) compared to the balance group.

Voxel to voxel functional connectivity across the entire connectome revealed significant end-point differences in patterns of functional connectivity of the anterior cingulate cortex (F=4.50, k=93, MNI = −04, +30, −04, size p-FWE = 0.02, size p-FDR= 0.02, peak p-FWE= 0.04) (Figure 1a), after controlling for pre-test functional connectivity and other covariates. Post hoc characterization of ACC functional connectivity patterns using seed to voxel analysis revealed that differences between interventions were driven by a mix of both more positive and more negative connectivity with the ACC in the aerobic compared to balance intervention (Figure 1b and 1C and Table 2). Individual trajectories of each ACC-to-cluster connection for aerobic and balance interventions are found in the supplementary materials.

**Figure 1:**
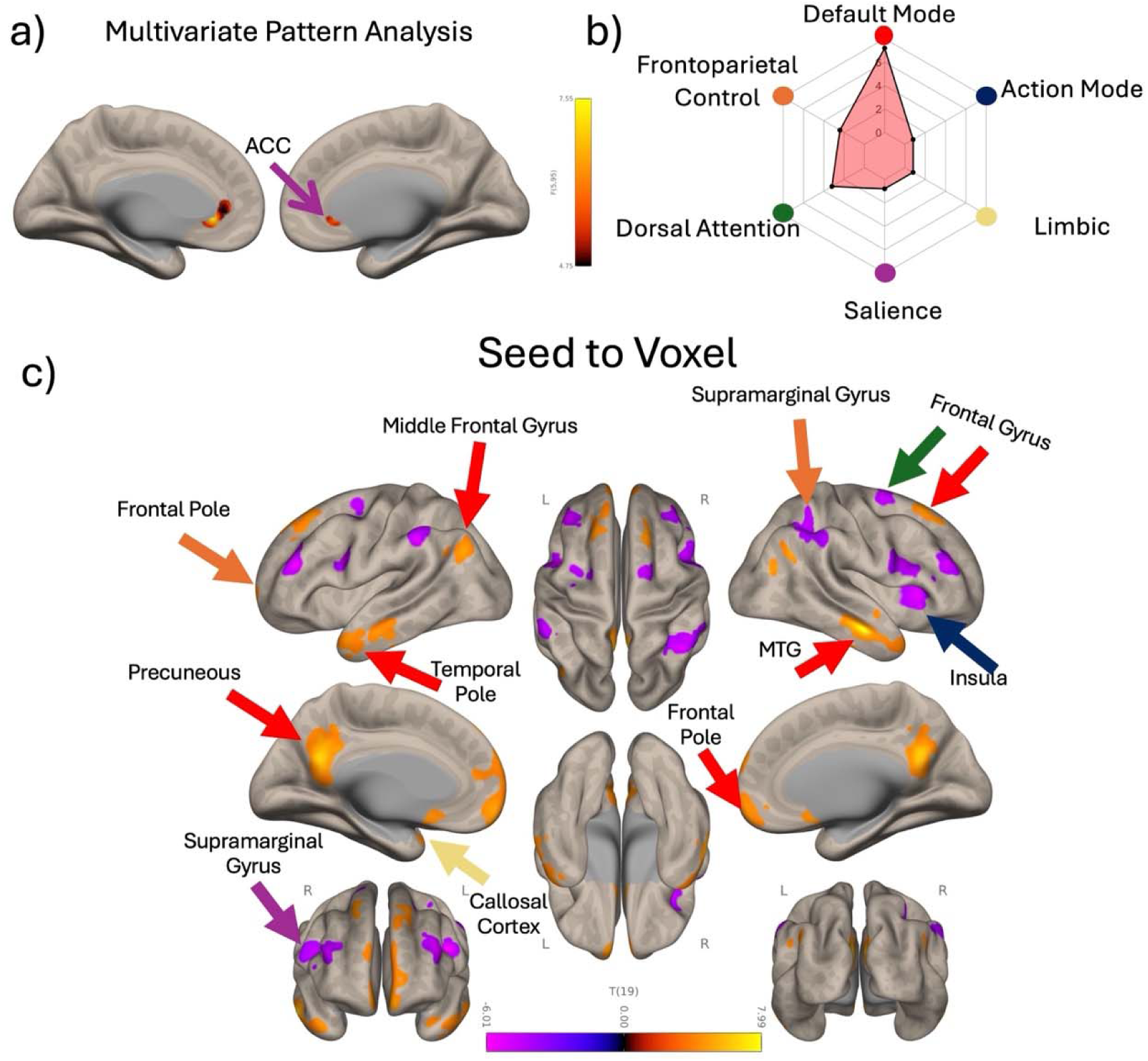
Intervention results from resting state fMRI. a) Voxel-to-voxel functional connectivity across the entire connectome revealed significant differences between aerobic and balance in the functional connectivity patterns of the anterior cingulate cortex. b) The radar plot provides a visual summary of how the significant post-hoc seed-to-voxel clusters are distributed across the seven intrinsic resting-state networks defined by Yeo et al.,(2011), where we label the ventral attention network as the newly defined Action Mode Network, with the x-axis representing each network name and the y-axis showing the number of significant voxels. Each cluster was assigned to a specific network based on its peak MNI coordinate, as determined using FMRIB’s Software Library (FSL). c) Post hoc seed-to-voxel results for the significant MVPA cluster revealed 19 clusters. Patterns of ACC functional connectivity differences in aerobic versus balance are driven by a mix of more positive and more negative connectivity within and between large-scale intrinsic functional networks, particularly within and between the DMN and FPN. Default Mode (red), Frontoparietal Control (orange), Dorsal Attention (Green), Salience (purple), Limbic (cream), Action Mode (Navy blue) (See supplementary Materials for expanded explanation). Orange/yellow clusters indicates hyperconnectivity with the ACC and purple/blue clusters indicates hypoconnectivity with the ACC.

**Table 2.** Post hoc full models. FDR, false discovery rate. FWE, family-wise error.

| **X** | **Y** | **Z** | **Size** | **Network** | **Region** | **Size p-FWE** | **Size p-FDR** | **Peak p-FWE** |
| --- | --- | --- | --- | --- | --- | --- | --- | --- |
| 8 | -52 | 20 | 1225 | Default Mode | Precuneus | <0.001 | <0.001 | 0.164416 |
| 68 | -4 | -18 | 886 | Default Mode | Medial temporal Gyrus | <0.001 | <0.001 | <0.001 |
| -4 | 62 | -18 | 661 | Default Mode | Frontal Pole | <0.001 | <0.001 | 0.645455 |
| 38 | -52 | 46 | 508 | Frontoparietal Control | Supramarginal gyrus | <0.001 | <0.001 | 0.614382 |
| 32 | 36 | 18 | 441 | Frontoparietal Control | Frontal pole | <0.001 | <0.001 | 0.111148 |
| -38 | 38 | 22 | 404 | Frontoparietal Control | Frontal pole | <0.001 | <0.001 | 0.107967 |
| 52 | 18 | 20 | 392 | Dorsal Attention | Inferior frontal gyrus | <0.001 | <0.001 | 0.722716 |
| -60 | -8 | -22 | 251 | Default Mode | Medial temporal gyrus | <0.01 | <0.001 | 0.908897 |
| -54 | -36 | 38 | 229 | Salience | Supramarginal Gyrus | <0.01 | <0.001 | 0.37766 |
| -42 | 6 | -30 | 223 | Default Mode | Temporal Pole | <0.01 | <0.001 | 0.874375 |
| -18 | 36 | 54 | 214 | Default Mode | Sup. Frontal Gyrus | 0.012174 | <0.001 | 0.606346 |
| 30 | 24 | 4 | 209 | Action Mode | Insula | 0.013644 | <0.001 | 0.688472 |
| -48 | -74 | 36 | 207 | Default Mode | Lateral Occipital | 0.014284 | <0.001 | 0.914627 |
| 22 | 32 | 58 | 194 | Default Mode | Sup. Frontal Gyrus | 0.019304 | <0.01 | 0.53122 |
| 0 | 12 | -12 | 163 | Limbic | Sub callosal cortex | 0.04055 | <0.01 | 0.548501 |
| -46 | 2 | 26 | 150 | Dorsal Attention | Precentral Gyrus | 0.055945 | 0.010518 | 0.078847 |
| 24 | 0 | 68 | 145 | Dorsal Attention | Sup. Frontal Gyrus | 0.063422 | 0.011266 | 0.807998 |
| 56 | -68 | 30 | 137 | Default Mode | Lateral Occipital | 0.077664 | 0.013129 | 0.088294 |
| -20 | -4 | 58 | 89 | Dorsal Attention | Sup. Frontal Gyrus | 0.270664 | 0.048556 | 0.751183 |

Correlation Between Change Differences in Functional Connectivity and Changes in Cognition Over Time Between groups: To test if changes in executive functioning over the intervention were a function of change differences in connectivity in the aerobic and balance group (extracting and testing the average of the ROIs in each network from the seed-to-voxel post hoc analysis), linear mixed-effects models revealed a significant group*time interaction with functional connectivity between the ACC and the Action Mode Network (insula) and TMT B scores (β =-68.55, SE=30.09, t=-2.278, 95% CI=(−131.76, −5.34), p=0.04) and TMT B-A scores (β = −86.43, SE=27.54, t=-3.138, 95% CI=(−144.29, −28.57), p=0.006). We also saw a significant interaction with functional connectivity between the ACC and Frontoparietal Control Network (Supramarginal gyrus and Frontal Pole) and TMT B-A scores (β= −89.726, SE=42.87, t=-2.09, 95% CI=(−173.7, −5.68), p=0.01). See table 3 for all results from all networks.

**Table 3:** Group*time with functional connectivity between the ACC and the networks and TMT scores.

| Network | Outcome | $\beta$ | SE | t | p | R<sup>2</sup> | Adj R<sup>2</sup> | 95% CI |
| --- | --- | --- | --- | --- | --- | --- | --- | --- |
| Action Mode | TMT-B change | -68.548 | 30.08 | -2.28 | 0.0351 | 0.51 | 0.32 | -127.5, -9.57 |
| Frontoparietal Control | TMT-B change | -89.726 | 42.87 | -2.09 | 0.0508 | 0.50 | 0.31 | -173.7, -5.68 |
| Salience | TMT-B change | -46.548 | 36.08 | -1.29 | 0.2134 | 0.45 | 0.24 | -117.2, 24.18 |
| Dorsal Attention | TMT-B change | -32.062 | 44.36 | -0.72 | 0.4792 | 0.39 | 0.16 | -119.0, 54.8959 |
| Limbic | TMT-B change | 12.5922 | 31.02 | 0.406 | 0.6896 | 0.39 | 0.15 | -48.20, 73.3918 |
| Default Mode | TMT-B change | -0.8424 | 34.79 | -0.02 | 0.9809 | 0.38 | 0.14 | -69.03, 67.3483 |
| Action Mode | TMT-B-A change | -86.43 | 27.54 | -3.14 | 0.0057 | 0.62 | 0.47 | -140.4, -32.45 |
| Frontoparietal Control | TMT-B-A change | -112.43 | 39.51 | -2.85 | 0.0107 | 0.60 | 0.45 | -189.8, -34.98 |
| Dorsal Attention | TMT-B-A change | -63.736 | 41.88 | -1.52 | 0.1455 | 0.50 | 0.30 | -145.8, 8.3623 |
| Salience | TMT-B-A change | -54.136 | 36.03 | -1.5 | 0.1503 | 0.49 | 0.29 | -124.7, 16.4914 |
| Limbic | TMT-B-A change | 39.8283 | 29.73 | 1.34 | 0.197 | 0.48 | 0.28 | -18.44, 98.10 |
| Default Mode | TMT-B-A change | 19.3315 | 34.79 | 0.556 | 0.5853 | 0.43 | 0.21 | -48.86, 87.52 |

*Post hoc* marginal effects analysis, taking the change in ACC and Action Mode Network connectivity and the change in TMT B-A at the group level revealed a significant positive association in the aerobic group (β = 46.92, SE=20.83, t=2.25, p=0.04) but a negative and non-significant association in the balance group (β =-39.51, SE=18.50, t=-2.14, p=0.05) (Figure 2), indicating that participants in the aerobic group who developed greater anticorrelation between the ACC and action mode network showed larger reductions in TMT B-A time. In TMT B, both groups were non-significant, with aerobic group having a positive association (β =37.76, SE=22.75, t=1.66, p=0.11) and balance having a negative association (β =-30.79, SE=20.21, t=-1.52, p=0.15). *Post hoc* marginal effects analysis for the change in ACC and frontoparietal control network connectivity and the change in TMT B at the group level revealed a significant negative association in the balance group (β = −58.18, SE=23.61, t=-2.46, p=0.02) but a negative and non-significant association in the aerobic group (β =54.24, SE=29.81, t=1.82, p=0.09)

**Figure 2:**
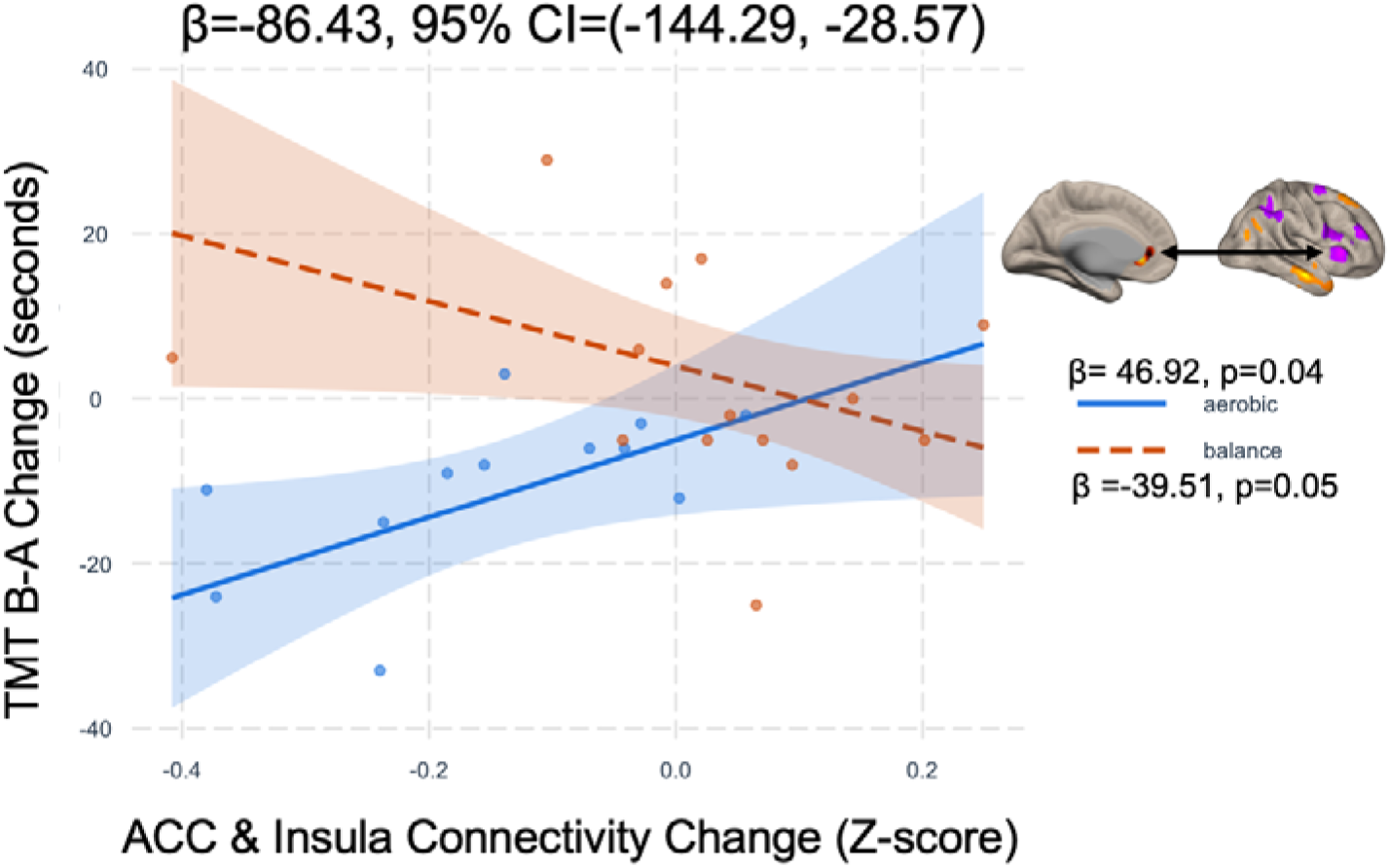
Line plot of the interaction between group and Anterior Cingulate Cortex (ACC) & Action Mode Network Connectivity Change. The relationship between ACC-Action Mode Network connectivity change and TMT B-A change differs significantly between the aerobic exercise and balance control groups. The aerobic group showed that the more anticorrelated the connection between the ACC and Action Mode Network, the smaller the completion time difference between TMT-B and TMT-A.

## Discussion

Using multivariate pattern analysis of resting-state fMRI, we found that 12 weeks of aerobic exercise, compared to active balance control, selectively modulated functional connectivity patterns of the anterior cingulate cortex (ACC), often involved in executive functioning. Results revealed widespread ACC connectivity changes across 19 cortical regions spanning default mode, frontoparietal control, and salience networks. Among these changes, the functional relationship between the ACC within the Salience Network and insula within the Action Mode Network emerged as the critical pathway mediating cognitive improvement: participants in the aerobic group who had greater ACC-insula anticorrelation showed significantly greater reductions in Trail Making Test B-A completion times. These findings suggest that aerobic exercise may enhance executive function after TBI through reorganization of ACC-insula connectivity, identifying this circuit as a possible therapeutic target for exercise-based rehabilitation interventions.

These current findings extend our prior cross-sectional evidence as well as prior interventional studies. Our prior cross sectional finding linked physical activity to the frontoparietal control network in TBI, where we showed associations between physical activity and functional connectivity of the inferior temporal gyrus (Tinney et al., 2024). While the earlier study established correlational relationships in TBI, the current randomized controlled trial provides preliminary causal evidence that aerobic exercise modulates functional connectivity, in a distinct pattern to that of non-aerobic balance exercises, after TBI. Additionally, prior interventional studies showed aerobic exercise improved functional neuroimaging outcomes in the ACC and thalamus in post-concussion syndrome(Leddy et al., 2012; Sharma et al., 2020). Our study extends these findings with a larger sample in community dwelling individuals. Notably, regions showing altered ACC connectivity in this intervention study overlap substantially with prior findings from España-Irla et al.’s characterization of post-TBI connectivity damage (España-Irla et al., 2025). Specifically, the frontal pole and the inferior frontal gyrus, which showed altered ACC connectivity between aerobic and balance groups, and was also identified in our prior cross-sectional physical activity analysis (Tinney et al., 2024). Additionally, there have been several reviews on functional connectivity in mTBI, where authors reported mTBI changes in functional connectivity of the default mode, frontoparietal, and salience network, three networks that are highly relevant in this current study(Dogra et al., 2024; Morelli et al., 2021), although there are inconsistencies with directionality. This suggests, perhaps, that aerobic exercise’s effect on functional brain patterns after TBI can modulate networks that themselves are damaged early after injury. The consistency of findings across these complementary lines of evidence helps to strengthens the hypothesis that exercise-induced connectivity changes in these regions represent neurobiological mechanisms underlying recovery, however caution should be taken given the pilot nature of this current study.

In the context of brain injury, the functional significance of connectivity patterns depends heavily on individual injury characteristics and recovery trajectories(Bickart et al., 2023; Bittencourt-Villalpando et al., 2021; Dogra et al., 2024; McDonald et al., 2012), making behavioral validation essential for interpretation(Bigler et al., 2025). In our main paper, we found that aerobic exercise improved TMT B and B-A performance(Tinney et al., 2025), so here we assessed if any of the connections between the ACC and resultant clusters correlated with the intervention effects on TMT. Specifically, we found that ACC-insula anticorrelation correlated with improved TMT performance differently in the aerobic group compared to the balance group. The differential correlation in the aerobic group argues against maladaptive disconnection (ACC-insula anticorrelation) but instead suggesting that exercise-induced functional segregation represents an adaptive mechanism enhancing cognitive flexibility. The spatial specificity is critical since the ACC and insula demonstrate strong positive connectivity in healthy individuals (Taylor et al., 2009), where the anterior insula serves as a salience detector generating representations of behaviorally relevant stimuli, with the ACC re-representing these signals to guide action selection(Medford & Critchley, 2010; Molnar-Szakacs & Uddin, 2022). The exercise-induced shift toward anticorrelation in rostral ACC and anterior insula may reflect adaptive decoupling between Salience Network affective regulatory circuits and Action Mode Network task-control circuits. While strong intrinsic connectivity facilitates rapid coordination between detection and control processes in healthy individuals(Badke D’Andrea et al., 2025; Taylor et al., 2009), functional segregation after brain injury may enable more dynamic, context-specific network configurations during task execution. Both the ACC(Bernacer et al., 2019; Li et al., 2017; Rosano et al., 2010; Yu et al., 2021) and insula(Albergoni et al., 2023; Schmitt et al., 2020; Williamson et al., 1999) demonstrate structural and functional plasticity in response to exercise across various populations, including post-concussion syndrome(Leddy et al., 2012).

Taken together, aerobic exercise may promote reorganization that preserves coordinated ACC-insula responses when executive demands require it, while potentially allowing greater functional independence during set-shifting tasks.

Importantly, different exercise modalities may produce cognitive benefits through distinct neural mechanisms. Our balance intervention, which emphasized controlled, deliberate movement and proprioceptive awareness, showed associations between ACC-frontoparietal segregation and improved TMT performance, while aerobic exercise demonstrated the ACC-insula relationship. This suggests that balance exercise may engage postural control and executive processes independently, benefiting from ACC-frontoparietal segregation(Kannan et al., 2023). In contrast, aerobic exercise, which engages cardiovascular and metabolic systems more directly, may drive cognitive improvements through different networks, as evidenced by the ACC-insula relationship. These preliminary observations suggest that balance and aerobic exercise may represent complementary, each targeting distinct aspects of brain network organization.

Several limitations in this study warrant consideration. First, the small sample size (n=24) limits statistical power and generalizability. However, the MVPA approach provides an advantage in low powered studies with increased sensitivity and reduced multiple comparisons(Nieto-Castanon, 2022). Second, sample heterogeneity, including injury timing (within one year), mechanisms, and recovery trajectories, reflects clinical reality but introduces variability.

Nevertheless, the data-driven approach possibly suggests a common outcome from aerobic exercise regardless of this heterogeneity. Larger samples however would enable injury-specific analyses and more definitive characterization of neural mechanisms. Third, differential sex representation across groups (92% female in aerobic vs. 42% in balance) is a limitation.

Evidence suggests sex differences in brain injury pathophysiology(Levin et al., 2021) and exercise responses(Ansdell et al., 2020; Parker et al., 2010), yet our sample precluded adequately powered stratified analyses. Future research should prioritize enrollment of more demographically diverse samples with adequate power for subgroup analyses. Additionally, the TMT represents only one measure of executive function and encompasses processing speed. The observed ACC-insula connectivity changes may be specific to TMT demands (set-switching, performance monitoring) rather than representing a general mechanism underlying all executive deficits. Nevertheless, TMT-B has been shown to be modulated with aerobic exercise in several TBI studies (Chin et al., 2015; López et al., 2024; Tinney et al., 2025). Future studies should examine whether exercise effects generalize to other executive measures and real-world functional outcomes such as return to work and independent living. Finally, optimal exercise parameters remain unclear. While our 12-week aerobic protocol modulated patterns of ACC connectivity in mild TBI, questions remain about efficacy in moderate-to-severe injuries, optimal intensity and duration, persistence of benefits after intervention cessation, and whether other exercise modalities produce similar effects. Future dose-response studies and direct comparisons between interventions are needed to optimize rehabilitation protocols.

## Conclusion

This study examined the neural mechanisms underlying aerobic exercise-induced cognitive improvements in individuals with mild TBI. The widespread pattern of ACC connectivity changes across default mode and frontoparietal networks suggests that aerobic exercise may facilitate network-level reorganization, relevant to mTBI-associated damage, and cognitive recovery. Specifically, the ACC-insula connectivity changes associate with TMT-B improvement following aerobic exercise intervention. These findings provide mechanistic evidence in support of aerobic exercise as a rehabilitative intervention for cognitive recovery following mTBI.

## Data Availability

Data will be available upon request.

## Acknowledgements

We would like to thank the participants for their participation in this study. We would also like to thank the TBI Lab undergraduate research assistants for their help in administering and monitoring the interventions. Additionally, this work was done in part with the Discovery and Explorer Cluster, supported by Northeastern University’s Research Computing team. The data was partially collected using REDCap, supported by NIH UL1TR002544.

## References

Albergoni, M., Storelli, L., Preziosa, P., Rocca, M. A., & Filippi, M. (2023). The insula modulates the effects of aerobic training on cardiovascular function and ambulation in multiple sclerosis. Journal of Neurology, 270(3), 1672–1681. 10.1007/s00415-022-11513-0

Ansdell, P., Thomas, K., Hicks, K. M., Hunter, S. K., Howatson, G., & Goodall, S. (2020). Physiological sex differences affect the integrative response to exercise: Acute and chronic implications. Experimental Physiology, 105(12), 2007–2021. 10.1113/EP088548

Badke D’Andrea, C., Laumann, T. O., Newbold, D. J., Lynch, C. J., Hadji, M., Nelson, S. M., Nielsen, A. N., Chauvin, R. J., Krimmel, S. R., Snyder, A. Z., Marek, S., Greene, D. J., Raichle, M. E., Dosenbach, N. U. F., & Gordon, E. M. (2025). Action-mode subnetworks for decision-making, action control, and feedback. Proceedings of the National Academy of Sciences, 122(27), e2502021122. 10.1073/pnas.2502021122

Behzadi, Y., Restom, K., Liau, J., & Liu, T. T. (2007). A component based noise correction method (CompCor) for BOLD and perfusion based fMRI. NeuroImage, 37(1), 90–101. 10.1016/j.neuroimage.2007.04.042

Bernacer, J., Martinez-Valbuena, I., Martinez, M., Pujol, N., Luis, E. O., Ramirez-Castillo, D., & Pastor, M. A. (2019). An amygdala-cingulate network underpins changes in effort-based decision making after a fitness program. NeuroImage, 203, 116181. 10.1016/j.neuroimage.2019.116181

Bickart, K., Sheridan, C., Frees, D., Kang, K., Fischer, J., Parks, C., & Kashou, A. (2023). A Systematic Review of Resting-State fMRI in Traumatic Brain Injury Across Injury Age, Severity, Mechanism, Chronicity, and Imaging Methods (P8-1.009). Neurology, 100(17_supplement_2), 4146. 10.1212/WNL.0000000000203779

Bigler, E. D., Allder, S., Dunkley, B. T., & Victoroff, J. (2025). What traditional neuropsychological assessment got wrong about mild traumatic brain injury. I: Historical perspective, contemporary neuroimaging overview and neuropathology update. Brain Injury, 39(14), 1161–1183. 10.1080/02699052.2025.2551162

Bittencourt-Villalpando, M., van der Horn, H. J., Maurits, N. M., & van der Naalt, J. (2021). Disentangling the effects of age and mild traumatic brain injury on brain network connectivity: A resting state fMRI study. NeuroImage: Clinical, 29, 102534. 10.1016/j.nicl.2020.102534

Chai, X. J., Castañón, A. N., Öngür, D., & Whitfield-Gabrieli, S. (2012). Anticorrelations in resting state networks without global signal regression. NeuroImage, 59(2), 1420–1428. 10.1016/j.neuroimage.2011.08.048

Chin, L. M., Keyser, R. E., Dsurney, J., & Chan, L. (2015). Improved Cognitive Performance Following Aerobic Exercise Training in People with Traumatic Brain Injury. Archives of Physical Medicine and Rehabilitation, 96(4), 754. 10.1016/J.APMR.2014.11.009

Cline, T. L., Morfini, F., Tinney, E., Makarewycz, E., Lloyd, K., Olafsson, V., Bauer, C. C. C., Kramer, A. F., Raine, L. B., Gabard-Durnam, L. J., Whitfield-Gabrieli, S., & Hillman, C. H. (2024). Resting-State Functional Connectivity Change in Frontoparietal and Default Mode Networks After Acute Exercise in Youth. Brain Plasticity, 9(1–2), 5–20. 10.3233/BPL-240003

Dogra, S., Arabshahi, S., Wei, J., Saidenberg, L., Kang, S. K., Chung, S., Laine, A., & Lui, Y. W. (2024). Functional Connectivity Changes on Resting-State fMRI after Mild Traumatic Brain Injury: A Systematic Review. American Journal of Neuroradiology, 45(6), 795–801. 10.3174/ajnr.A8204

Dosenbach, N. U. F., Raichle, M. E., & Gordon, E. M. (2025). The brain’s action-mode network. Nature Reviews Neuroscience, 26(3), 158–168. 10.1038/s41583-024-00895-x

Dupuy, O., Ludyga, S., Ortega, F. B., Hillman, C. H., Erickson, K. I., Herold, F., Kamijo, K., Wang, C.-H., Morris, T. P., Brown, B., Esteban-Cornejo, I., Solis-Urra, P., Bosquet, L., Gerber, M., Mekari, S., Berryman, N., Bherer, L., Rattray, B., Liu-Ambrose, T., … Cheval, B. (2024). Do not underestimate the cognitive benefits of exercise. Nature Human Behaviour, 8(8), 1460–1463. 10.1038/s41562-024-01949-x

España-Irla, G., Tinney, E. M., Ai, M., Nwakamma, M., & Morris, T. P. (2025). Functional Connectivity Patterns Following Mild Traumatic Brain Injury and the Association With Longitudinal Cognitive Function. Human Brain Mapping, 46(8), e70237. 10.1002/hbm.70237

Esteban, O., Markiewicz, C. J., Blair, R. W., Moodie, C. A., Isik, A. I., Erramuzpe, A., Kent, J. D., Goncalves, M., DuPre, E., Snyder, M., Oya, H., Ghosh, S. S., Wright, J., Durnez, J., Poldrack, R. A., & Gorgolewski, K. J. (2019). fMRIPrep: A robust preprocessing pipeline for functional MRI. Nature Methods, 16(1), 111–116. 10.1038/s41592-018-0235-4

Friston, K. J., Williams, S., Howard, R., Frackowiak, R. S. J., & Turner, R. (1996). Movement-Related effects in fMRI time-series. Magnetic Resonance in Medicine, 35(3), 346–355. 10.1002/mrm.1910350312

Hallquist, M. N., Hwang, K., & Luna, B. (2013). The nuisance of nuisance regression: Spectral misspecification in a common approach to resting-state fMRI preprocessing reintroduces noise and obscures functional connectivity. NeuroImage, 82, 208–225. 10.1016/j.neuroimage.2013.05.116

Ho, N. F., Han, S. P., & Dawe, G. S. (2009). Effect of voluntary running on adult hippocampal neurogenesis in cholinergic lesioned mice. BMC Neuroscience, 10(1), 57. 10.1186/1471-2202-10-57

Kannan, L., Bhatt, T., & Ajilore, O. (2023). Cerebello-cortical functional connectivity may regulate reactive balance control in older adults with mild cognitive impairment. Frontiers in Neurology, 14. 10.3389/fneur.2023.1041434

Leddy, J., Cox, J., Baker, J., Wack, D., & Willer, B. (2012). Exercise Treatment For Post Concussion Syndrome: A Placebo Controlled Pilot Study Of Changes In fMRIBlood Flow And Symptoms. Brain Injury, 26(4–5), 388–389.

Levin, H. S., Temkin, N. R., Barber, J., Nelson, L. D., Robertson, C., Brennan, J., Stein, M. B., Yue, J. K., Giacino, J. T., McCrea, M. A., Diaz-Arrastia, R., Mukherjee, P., Okonkwo, D. O., Boase, K., Markowitz, A. J., Bodien, Y., Taylor, S., Vassar, M. J., Manley, G. T., … Zafonte, R. (2021). Association of Sex and Age With Mild Traumatic Brain Injury-Related Symptoms: A TRACK-TBI Study. JAMA Network Open, 4(4), e213046. 10.1001/jamanetworkopen.2021.3046

Li, M., Huang, M., Li, S., Tao, J., Zheng, G., & Chen, L. (2017). The effects of aerobic exercise on the structure and function of DMN-related brain regions: A systematic review. International Journal of Neuroscience, 127(7), 634–649. 10.1080/00207454.2016.1212855

Lloyd, K. M., Morris, T. P., Anteraper, S., Voss, M., Nieto-Castanon, A., Whitfield-Gabrieli, S., Fanning, J., Gothe, N., Salerno, E. A., Erickson, K. I., Hillman, C. H., McAuley, E., & Kramer, A. F. (2024). Data-driven MRI analysis reveals fitness-related functional change in default mode network and cognition following an exercise intervention. Psychophysiology, 61(4), e14469. 10.1111/psyp.14469

López, L. P., Coll-Andreu, M., Torras-Garcia, M., Font-Farré, M., Oviedo, G. R., Capdevila, L., Guerra-Balic, M., Portell-Cortés, I., Costa-Miserachs, D., & Morris, T. P. (2024). Aerobic exercise and cognitive function in chronic severe traumatic brain injury survivors: A within-subject A-B-A intervention study. BMC Sports Science, Medicine and Rehabilitation, 16(1), 201. 10.1186/s13102-024-00993-4

McDonald, B. C., Saykin, A. J., & McAllister, T. W. (2012). Functional MRI of mild traumatic brain injury (mTBI): Progress and perspectives from the first decade of studies. Brain Imaging and Behavior, 6(2), 193–207. 10.1007/s11682-012-9173-4

Medford, N., & Critchley, H. D. (2010). Conjoint activity of anterior insular and anterior cingulate cortex: Awareness and response. Brain Structure and Function, 214(5), 535–549. 10.1007/s00429-010-0265-x

Mishra, A., Hall, C. N., Howarth, C., & Freeman, R. D. (2020). Key relationships between non-invasive functional neuroimaging and the underlying neuronal activity. Philosophical Transactions of the Royal Society B: Biological Sciences, 376(1815), 20190622. 10.1098/rstb.2019.0622

Molnar-Szakacs, I., & Uddin, L. Q. (2022). Anterior insula as a gatekeeper of executive control. Neuroscience & Biobehavioral Reviews, 139, 104736. 10.1016/j.neubiorev.2022.104736

Moore, D., Jung, M., Hillman, C. H., Kang, M., & Loprinzi, P. D. (2022). Interrelationships between exercise, functional connectivity, and cognition among healthy adults: A systematic review. Psychophysiology, 59(6), e14014. 10.1111/psyp.14014

Morelli, N., Johnson, N. F., Kaiser, K., Andreatta, R. D., Heebner, N. R., & Hoch, M. C. (2021). Resting state functional connectivity responses post-mild traumatic brain injury: A systematic review. Brain Injury, 35(11), 1326–1337. 10.1080/02699052.2021.1972339

Morris, T., Gomes Osman, J., Tormos Muñoz, J. M., Costa Miserachs, D., & Pascual Leone, A. (2016). The role of physical exercise in cognitive recovery after traumatic brain injury: A systematic review. Restorative Neurology and Neuroscience, 34(6), 977–988. 10.3233/RNN-160687

Nieto-Castanon, A. (2022). Brain-wide connectome inferences using functional connectivity MultiVariate Pattern Analyses (fc-MVPA). PLoS Computational Biology, 18(11), e1010634. 10.1371/journal.pcbi.1010634

Parker, B. A., Kalasky, M. J., & Proctor, D. N. (2010). Evidence for sex differences in cardiovascular aging and adaptive responses to physical activity. European Journal of Applied Physiology, 110(2), 235–246. 10.1007/s00421-010-1506-7

Penny, W. D., Friston, K. J., Ashburner, J. T., Kiebel, S. J., & Nichols, T. E. (2011). Statistical Parametric Mapping: The Analysis of Functional Brain Images. Elsevier.

Piao, C.-S., Stoica, B. A., Wu, J., Sabirzhanov, B., Zhao, Z., Cabatbat, R., Loane, D. J., & Faden, A. I. (2013). Late exercise reduces neuroinflammation and cognitive dysfunction after traumatic brain injury. Neurobiology of Disease, 54, 252–263. 10.1016/j.nbd.2012.12.017

Power, J. D., Mitra, A., Laumann, T. O., Snyder, A. Z., Schlaggar, B. L., & Petersen, S. E. (2014). Methods to detect, characterize, and remove motion artifact in resting state fMRI. NeuroImage, 84, 320–341. 10.1016/j.neuroimage.2013.08.048

Rabinowitz, A. R., & Levin, H. S. (2014). Cognitive Sequelae of Traumatic Brain Injury. Psychiatric Clinics, 37(1), 1–11. 10.1016/j.psc.2013.11.004

Rosano, C., Venkatraman, V. K., Guralnik, J., Newman, A. B., Glynn, N. W., Launer, L., Taylor, C. A., Williamson, J., Studenski, S., Pahor, M., & Aizenstein, H. (2010). Psychomotor Speed and Functional Brain MRI 2 Years After Completing a Physical Activity Treatment. The Journals of Gerontology: Series A, *65A*(6), 639–647. 10.1093/gerona/glq038

Schmitt, A., Upadhyay, N., Martin, J. A., Rojas Vega, S., Strüder, H. K., & Boecker, H. (2020). Affective Modulation after High-Intensity Exercise Is Associated with Prolonged Amygdalar-Insular Functional Connectivity Increase. Neural Plasticity, 2020(1), 7905387. 10.1155/2020/7905387

Sharma, B., Allison, D., Tucker, P., Mabbott, D., & Timmons, B. W. (2020). Cognitive and neural effects of exercise following traumatic brain injury: A systematic review of randomized and controlled clinical trials. Brain Injury, 34(2), 149–159. 10.1080/02699052.2019.1683892

Snowden, T., Morrison, J., Boerstra, M., Eyolfson, E., Acosta, C., Grafe, E., Reid, H., Brand, J., Galati, M., Gargaro, J., & Christie, B. R. (2023). Brain changes: Aerobic exercise for traumatic brain injury rehabilitation. Frontiers in Human Neuroscience, 17. 10.3389/fnhum.2023.1307507

Taylor, K. S., Seminowicz, D. A., & Davis, K. D. (2009). Two systems of resting state connectivity between the insula and cingulate cortex. Human Brain Mapping, 30(9), 2731–2745. 10.1002/hbm.20705

Thomas Yeo, B. T., Krienen, F. M., Sepulcre, J., Sabuncu, M. R., Lashkari, D., Hollinshead, M., Roffman, J. L., Smoller, J. W., Zöllei, L., Polimeni, J. R., Fischl, B., Liu, H., & Buckner, R. L. (2011). The organization of the human cerebral cortex estimated by intrinsic functional connectivity. Journal of Neurophysiology, 106(3), 1125–1165. 10.1152/jn.00338.2011

Tinney, E. M., Ai, M., España-Irla, G., Hillman, C. H., & Morris, T. P. (2024). Physical activity and frontoparietal network connectivity in traumatic brain injury. Brain and Behavior, 14(9), e70022. 10.1002/brb3.70022

Tinney, E. M., Nwakamma, M. C., España-Irla, G., Kong, L., Chen, C., Hwang, J., O’Brien, A., Perko, M., Sodemann, R. L., Caefer, J., Manczurowsky, J., Stillman, A., Hillman, C. H., & Morris, T. P. (2025). The feasibility and efficacy of a virtual, symptom-guided aerobic exercise intervention to improve cognition in mild traumatic brain injury: A single-blind pilot randomized control trial with an active comparator group. medRxiv, 2025.12.11.25342088. 10.64898/2025.12.11.25342088

Tinney, E. M., Nwakamma, M. C., España-Irla, G., Perko, M., Sodemann, R. L., Caefer, J., Manczurowsky, J., Hillman, C. H., Stillman, A., & Morris, T. P. (2026). The Exercise and Concussion Health Study (TECHS): Pilot and Feasibility Protocol. Contemporary Clinical Trials Communications, 101608. 10.1016/j.conctc.2026.101608

Voss, M. W., Prakash, R. S., Erickson, K. I., Basak, C., Chaddock, L., Kim, J. S., Alves, H., Heo, S., Szabo, A., White, S. M., Wojcicki, T. R., Mailey, E. L., Gothe, N., Olson, E. A., McAuley, E., & Kramer, A. F. (2010). Plasticity of Brain Networks in a Randomized Intervention Trial of Exercise Training in Older Adults. Frontiers in Aging Neuroscience, 2. 10.3389/fnagi.2010.00032

Whitfield-Gabrieli, S., & Nieto-Castanon, A. (2012). Conn: A Functional Connectivity Toolbox for Correlated and Anticorrelated Brain Networks. Brain Connectivity, 2(3), 125–141. 10.1089/brain.2012.0073

Williamson, J. W., McColl, R., Mathews, D., Ginsburg, M., & Mitchell, J. H. (1999). Activation of the insular cortex is affected by the intensity of exercise. Journal of Applied Physiology, 87(3), 1213–1219. 10.1152/jappl.1999.87.3.1213

Worsley, K. J., Marrett, S., Neelin, P., Vandal, A. C., Friston, K. J., & Evans, A. C. (1996). A unified statistical approach for determining significant signals in images of cerebral activation. Human Brain Mapping, 4(1), 58–73. 10.1002/(SICI)1097-0193(1996)4:1%253C58::AID-HBM4%253E3.0.CO;2-O

Yu, Q., Herold, F., Becker, B., Klugah-Brown, B., Zhang, Y., Perrey, S., Veronese, N., Müller, N. G., Kramer, A. F., & Zou, L. (2021). Cognitive benefits of exercise interventions: An fMRI activation likelihood estimation meta-analysis. Brain Structure and Function, 226(3), 601–619. 10.1007/s00429-021-02247-2

Zhang, P., Yu, H., Zhou, N., Zhang, J., Wu, Y., Zhang, Y., Bai, Y., Jia, J., Zhang, Q., Tian, S., Wu, J., & Hu, Y. (2013). Early exercise improves cerebral blood flow through increased angiogenesis in experimental stroke rat model. Journal of NeuroEngineering and Rehabilitation, 10(1), 43. 10.1186/1743-0003-10-43

